# Effectiveness of mass dengue vaccination in the state of Paraná, Brazil: integrating case-cohort and case-control designs

**DOI:** 10.1101/2023.08.29.23292476

**Authors:** Fredi Alexander Diaz-Quijano, Denise Siqueira de Carvalho, Sonia Mara Raboni, Silvia Emiko Shimakura, Angela Maron de Mello, Magda Clara Vieira da Costa-Ribeiro, Lineu Silva, Marilene da Cruz Magalhães Buffon, Eliane Mara Cesario Pereira Maluf, Gabriel Graeff, Gustavo Almeida, Clara Preto, Karin Regina Luhm

**Affiliations:** Department of Epidemiology, Laboratory of Causal Inference in Epidemiology – LINCE-USP, School of Public Health, University of São Paulo, São Paulo, SP, 01246-904, Brazil; Department of Public Health, Federal University of Paraná, Curitiba, Brazil; Department of Statistics, Federal University of Paraná, Curitiba, Brazil; Postgraduate program in Public Health, Federal University of Paraná, Curitiba, Brazil; Health Ministry, Curitiba, Brazil; Department of Basic Pathology and postgraduate program in Microbiology, Parasitology and Pathology, Federal University of Paraná, Curitiba, Brazil; Foundation of the Federal University of Paraná, Curitiba, Brazil

## Abstract

We aimed to estimate the effectiveness of CYD-TDV in preventing symptomatic dengue cases during a campaign targeting individuals aged 15-27 years in selected municipalities in Paraná, Brazil. Additionally, we examined whether a history of dengue, as recorded by the surveillance system, modified the vaccine’s effectiveness.

**Methods:** We conducted a case-cohort analysis comparing the frequency of vaccination, with at least one dose of CYD-TDV, in individuals confirmed to have dengue by RT-PCR, identified by the surveillance system during 2019 and 2020, with the vaccination coverage in the target population. Moreover, with a case-control design using weighted controls, we assessed the history of dengue as a modifier of the vaccine’s effectiveness. The analyses were performed using a logistic random-effects regression model, with data clustered in municipalities and incorporating covariates such as the incidence of dengue before the campaign, age, and sex.

**Results:** During the study period, 1,869 cases of dengue were identified. The vaccination frequency among these cases was significantly lower than the overall vaccination coverage of the participating municipalities (50.4% vs. 57.2%, respectively; adjusted odds ratio: 0.79; 95% confidence interval: 0.72–0.87). In individuals with a history of dengue, vaccination was more than 70% effective in reducing the incidence of dengue. However, vaccination was not associated with significantly reducing the overall dengue case risk in individuals without a history of dengue.

**Conclusion:** Vaccination significantly decreased dengue cases in the target population. The case-control design suggested that this reduction was primarily driven by the benefits seen in individuals with a history of dengue. Previous dengue diagnosis recorded by epidemiological surveillance could serve as a criterion for the recommendation of CYD-TDV, especially in endemic regions with limited serological testing facilities.

## INTRODUCTION

Dengue viruses are one of the most significant arboviruses worldwide, and it is estimated that nearly half of the global population is at risk of infection [1]. As no specific treatment is available, intervention efforts for the disease have primarily focused on controlling the vectors that transmit the virus [2]. CYD-TDV (Dengvaxia®) was the first vaccine for dengue to receive approval and was launched in Brazil in 2015 for individuals aged 9-44 years and administered in three doses with an interval of six months between them [3,4]. It was subsequently recommended by the World Health Organization (WHO) in 2016 for use in regions with a high burden of the disease [3].

In Southern Brazil, the state of Paraná has experienced outbreaks of dengue since the 1990s. The incidence rate reached 462 cases per 100,000 inhabitants from August 2015 to July 2016 [5]. In response, the government of Paraná offered the dengue vaccine from August 2016 to December 2018 [5] to a target population of 500,000 individuals in 30 municipalities due to the high incidence of the disease, predominantly in the 15 to 27 age group. During the campaign, 302,603 people were vaccinated, with vaccination coverage of 60.5% for at least one dose, 44.2% for two or more doses, and 28.6% for three doses [6].

In 2017, a re-evaluation of clinical trials suggested that the vaccine was only effective in protecting individuals previously infected with dengue [7]. This new information might have negatively impacted adherence to the full vaccination course. In 2018, the WHO also recommended that the vaccine only is given to individuals with a history of laboratory-confirmed dengue infection or regions with a prevalence > 80% [8]. However, this recommendation may create a barrier to access as diagnostic tests to confirm past infection are not widely available. Furthermore, it raises questions about the overall benefit of the mass vaccination campaign and whether using the tools available in surveillance to identify individuals with a history of dengue can aid in identifying those who would benefit the most from the vaccine. In addition, the cross-reactivity between antibody responses to Zika and dengue viruses may further complicate the identification of seropositive individuals in regions where both viruses co-circulate [9].

The present study aimed to estimate the effectiveness of the CYD-TDV vaccine on the incidence of dengue in the individuals vaccinated during the campaign in Paraná. Additionally, since access to serological tests was limited, we examined whether a history of dengue, as recorded by the surveillance system, modified the vaccine’s effectiveness.

## METHODS

### Study design and population

We conducted a case-cohort analysis comparing the frequency of vaccination, with at least one dose of CYD-TDV, in individuals with dengue confirmed by the surveillance system and the vaccination coverage in the target population. Additionally, a case-control design was used to assess whether a history of dengue modified the vaccine’s effectiveness.

The state of Paraná, located in the southern region of Brazil, had 399 municipalities and an estimated population of 11,242,720 inhabitants in 2016. The vaccination campaign targeted individuals aged 15 to 27 years in 28 municipalities with frequent epidemics. Two other municipalities with an incidence greater than 8,000 cases per 100,000 inhabitants in the preceding year of campaign, vaccinated individuals from 9 to 44 [6]. For this study, we included only the municipalities that, besides participating in the campaign, reported dengue cases during 2019 and 2020. Furthermore, we limited the study population to individuals 15-27 years old during the campaign, which was the common age group among all the included municipalities.

We defined cases of dengue as those confirmed by RT-PCR, as reported on Jan 1, 2019, to Dec 31, 2020 (when the last doses of the campaign had been applied more than 30 days ago) in the Notifiable Diseases Information System (*Sistema de Informação de Agravos de Notificação* - Sinan), a national surveillance system, or in the Local Environment Manager (*Gerenciador de Ambiente Local* - GAL), an information system for public health laboratories. Secondary outcomes included serotype-specific cases and dengue hospitalizations.

In the case-cohort design, we defined a population cohort as a group of individuals aged 15-27 years according to the Brazilian Institute of Geography and Statistics (*Instituto Brasileiro de Geografia e Estatística* - IBGE) projections for each age group, sex, and participating municipality [10]. The proportion of the exposed cohort for each category was calculated using vaccination records maintained by the government during the campaign [6].

In the case-control design, two groups of controls were chosen to assess the consistency of the associations [11]. The first group consisted of individuals suspected of having dengue but tested negative (TN) by RT-PCR and other confirmatory tests, such as NS1 antigen and IgM antibody. The second group consisted of individuals reported on Sinan as having other health problems (OHP), including reportable diseases unrelated to dengue: attendance for anti-rabies prophylaxis, exogenous poisoning, and accidents with venomous animals. Cases and controls living in peri-urban or rural areas were excluded from the analysis because they might have limited access to healthcare facilities compared to urban populations, which might have resulted in an increased likelihood of underreporting dengue history. The data obtained from IBGE indicated that 4.3% of the people in the 28 municipalities included in the study lived in rural areas, and only 2.8% of reported dengue cases originated from peri-urban or rural areas.

### Data sources

All municipalities participating in the campaign recorded data on vaccinated individuals in a nominal computerized database that included identification (name, sex, and date of birth) and information on administered vaccine doses.

All suspected cases of reportable diseases in the country, treated in the public or private health system, are reported on Sinan, the source for identifying cases and controls. This system includes demographic, clinical, and laboratory data. Suspected dengue cases were defined as those individuals who lived in or had travelled to an area where dengue transmission was active or the vector was present and who had fever and at least two of the following symptoms: nausea/vomiting, rash, muscle/joint pain, headache/pain behind the eyes, petechiae/positive tourniquet test result, and low white blood cell count. The surveillance system subsequently classified the reported case based on information from laboratory and clinical-epidemiological tests [12].

Case and control information was linked to the individual vaccination database using probabilistic procedures with the OpenRecklink software, version 3.1.824.4086. Independent reviewers implemented seven blocking strategies (Supplementary material, Table S1). The criteria for inclusion as a possible pair in the blocking were: concordance of the patient’s name and mother’s name (above 0.01), year of birth (above 0.74), sex, and municipality (exact). At least two individuals conducted the procedure, and the authors reviewed any disagreements.

### Statistical analysis

In the case-cohort design, a database was created to represent the target population in all participating municipalities that had reported dengue cases during the observation period. The database was used to recreate the distribution of inhabitants according to vaccination status, sex, and two age groups (15-20 and 21-27 years) in each of the municipalities. According to the campaign database, these values were calculated using the population estimates from IBGE and the count of vaccinated individuals in each pattern of covariates.

The population cohort information did not distinguish between urban, peri-urban, and rural areas because the vaccine database did not include information on the location of residence. However, we observed that the vaccination frequency of both cases and controls in the urban region was similar to those excluded from the rural and peri-urban areas (cases: 50.3% vs. 46.3%, p=0.58; TN controls: 10.4% vs. 6%, p=0.18, OHP controls: 48.8% vs. 48.9%, p=0.97). Based on this, we assumed that the analysed vaccination coverage distribution represents the urban population from which the cases proceeded.

We pre-selected the adjustment variables through a directed acyclic diagram to estimate vaccine effectiveness. Previous incidence of dengue is thought to affect ‘individuals’ perception of risk, which can subsequently influence their knowledge, attitudes, and practices, including their interest in vaccinating [13,14]. Additionally, previous incidence can affect the subsequent risk of infection, along with other contextual factors, including the degree of urbanization and any control actions adopted by the municipality. Furthermore, age and sex can affect campaign adherence and the risk of transmission and disease, and age can be a determinant of previous dengue infections in endemic areas (Figure 1).

**Figure 1.**
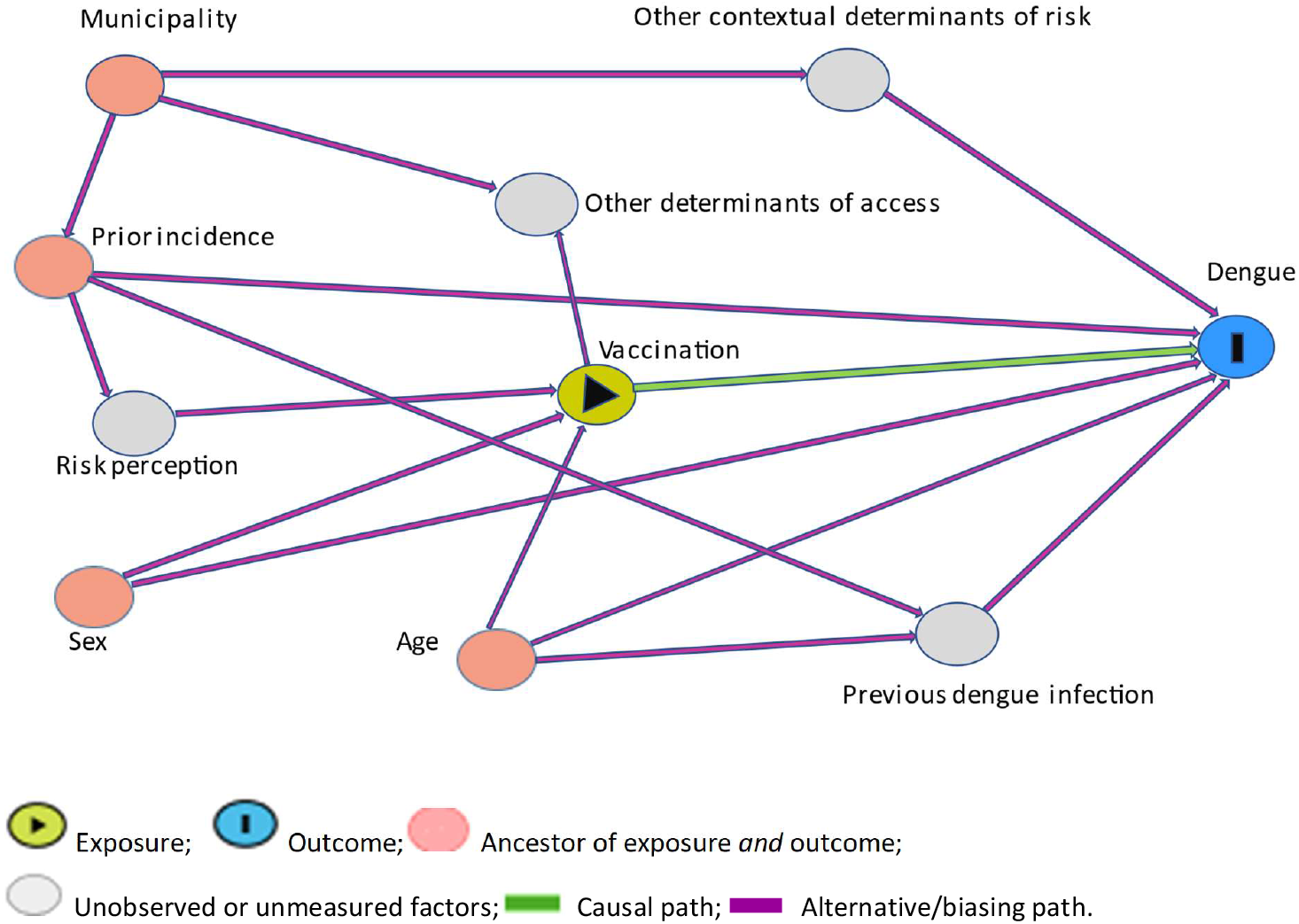
Directed acyclic diagram (DAG) representing the causal relationship between vaccine, dengue, and other covariates.

To estimate the effectiveness, we used a random effects logistic regression model with data aggregated at the municipal level by adjusting for the incidence of dengue in the municipality of residence in the year before the campaign (August 1, 2015-July 30, 2016), as a contextual determinant, and age and sex, as individual covariates. In this way, we compared the vaccination frequency between cases and the target population cohort.

The case-control design used weighting to ensure the controls accurately reflected the target population’s characteristics. The weight assigned to each control corresponded to the inverse probability of being included in the study based on vaccination status, sex, age group (15-20 and 21-27 years), and area of residence. Aiming to avoid having too few controls according to covariate patterns, municipalities were grouped into four areas based on proximity (Supplementary materials: Figure S1, Table S2). The estimates were obtained from random effects logistic models that, in addition to the prior municipality incidence (contextual), included age as a continuous variable, sex, history of dengue, and an interaction term between vaccination and history of dengue. We defined the history of dengue as a record of a probable case of dengue reported on Sinan, with the onset of symptoms between January 2008 and July 2016. The surveillance system defined probable cases as those confirmed by laboratory criteria, those classified by clinical-epidemiological criteria, or inconclusive cases. Besides at least one dose, with the case-control design, we also estimated the effectiveness of three doses of the vaccine as compared with no vaccination.

As both cohort and weighed controls were intended to represent the baseline vaccination coverage in the source population, all the odds ratios (ORs) reported here are interpretable as relative risk estimates [15–18]. Therefore, we calculated vaccine effectiveness (VE) as (1-OR) x 100%. The statistical analyses were performed using STATA 17.0 (StataCorp, College Station, Texas, USA).

## RESULTS

### Case-cohort design

From Jan 1, 2019, to Dec 31, 2020, 1,869 dengue cases were identified in individuals residing in urban areas aged 15 to 27 years in 28 municipalities that participated in the campaign. When comparing the cases with the target population in these municipalities, we observed a similar distribution of age groups but a slight difference in the distribution of sex. The likelihood of being vaccinated for dengue cases was significantly lower than that of the population (50.4% vs. 57.2%, respectively) (Table 1), as evidenced by a crude OR of 0.76 (95% confidence interval [CI]: 0.69–0.83; p<0.001). This measure was 0.79 (95% CI: 0.72–0.87) when adjusted for sex, age group, and previous incidence of dengue in a multilevel model (Table 2).

**Table 1.** General description of dengue cases and the target population for vaccination, 28 municipalities, Paraná, 2019 -2020.

| Variable | Cases (n=1,869) | Population cohort (n= 441,945)* |
| --- | --- | --- |
| Sex (male) – n (%) | 869 (46.5) | 219,589 (49.7) |
| Age group |  |  |
| 15–20 years old | 876 (46.9) | 202,076 (45.7) |
| 21–27 years old | 993 (53.1) | 239,869 (54.3) |
| Vaccinated | 941 (50.4) | 252,820 (57.2) |

\* Estimated population of 28 municipalities participating in the campaign that reported cases of dengue from 2019 to 2020.

**Table 2.** Pooled effect of vaccination with at least one dose of CYD-TDV on dengue outcomes, in 28 municipalities of Paraná, 2019 -2020.

| Outcome variables | Number of cases | OR (95% CI) * | VE – % (95% CI) | p-value |
| --- | --- | --- | --- | --- |
| Total dengue cases | 1,869 | 0.79 (0.72 – 0.87) | 21.3 (13.4 – 28.4) | <0.001 |
| Serotype-specific cases† |  |  |  |  |
| DENV-1 | 576 | 0.57 (0.48 – 0.68) | 42.7 (31.9 – 51.7) | <0.001 |
| DENV-2 | 1,162 | 1.08 (0.95 – 1.22) | -7.8 (-21.8 – 4.7) | 0.23 |
| DENV-4 ‡ | 130 | 0.13 (0.07 – 0.23) | 87.2 (77.2 – 92.8) | <0.001 |
| Hospitalisations | 100 | 1.12 (0.74 – 1.7) | -12.3 (-70 – 25.8) | 0.58 |

\* ORs adjusted for previous municipal incidence, age, and sex in a logistic random effects model with data aggregated at the municipal level.
† No cases of DENV-3 were identified in the study.
‡ Serotype 4 was analysed in only two neighbouring municipalities which included 130 of the 131 cases of DENV-4. Therefore, no additional conditioning for municipal variables was applied.

Vaccination was associated with a significant reduction in the incidence of dengue caused by serotype 1. However, no significant association was found between vaccination and cases caused by serotype 2. For serotype 4, we restricted the statistical analysis to two neighbouring municipalities, Foz do Iguaçu and Santa Terezinha do Itaipu, where 130 out of 131 cases were identified. Vaccination was associated with an 87% decrease in the incidence of DENV-4 (OR: 0.13; 95% CI: 0.07–0.23). No cases of serotype 3 were identified during the study period. There was no significant association between vaccination and the total number of hospitalizations for dengue (Table 2).

### Case-control design to assess the vaccine effectiveness modification by reported dengue history

Compared with the cases, both the control groups had similar distributions regarding demographic variables (Table 3). We observed that the unweighted estimates of the association between vaccination and dengue were positive (OR>1). However, weighted estimates were similar to those obtained in the case-cohort design, especially when comparing cases to the OHP control group. The association measures did not change substantially when adjusting for age (available as a quantitative variable only for the case-control design). Furthermore, there was no significant change in the OR value when adjusting for the previous incidence of dengue (Supplementary material: Table S3).

**Table 3.** Baseline characteristics of cases and controls, 28 municipalities, Paraná, 2019 -2020.

| Variable | Cases<br>(n=1869) | TN controls<br>(n=3897) | OHP controls<br>(n=5595) |
| --- | --- | --- | --- |
| Sex (male) – n (%) | 869 (46.5) | 1781 (45.7) | 2488 (44.7) |
| Age (years)* | 23 (20 – 27) | 24 (21 – 27) | 23 (20 – 27) |
| Pregnant (female population)(%) † | 61 (6.1) | 149 (7.0) | 120 (3.9) |
| Race (%) |  |  |  |
| White | 1403(75.0) | 2924(75.0) | 4159(74.7) |
| Brown | 287(15.4) | 607(15.6) | 245(4.4) |
| Black | 85(4.6) | 164(4.2) | 46(0.8) |
| Asian | 18(1.0) | 32(0.8) | 874(15.7) |
| Indigenous | 5(0.3) | 3(0.1) | 18(0.3) |
| Not informed | 71(3.8) | 167(4.3) | 223(4.1) |
| Education level (%) |  |  |  |
| Incomplete primary (<4<sup>th</sup> grade) | 22(1.2) | 31(0.8) | 59(1.1) |
| Incomplete primary (≥4<sup>th</sup> to <8<sup>th</sup> grade) | 157(8.4) | 309(7.9) | 455(8.2) |
| Complete primary | 325(17.4) | 698(17.9) | 1074(19.3) |
| Complete secondary | 680(36.4) | 1352(34.7) | 2539(45.6) |
| Higher education | 130(7.0) | 235(6.0) | 486(8.7) |
| Not informed | 555(29.6) | 1272(32.7) | 952(17.1) |
| Vaccination | 22(1.2) | 31(0.8) | 59(1.1) |
| No. of vaccine doses administered (%) |  |  |  |
| None | 928 (49.7) | 3492 (89.6) | 2847 (51.2) |
| 1 | 231 (12.4) | 117 (3.0) | 654 (11.8) |
| 2 | 256 (13.7) | 104 (2.7) | 729 (13.1) |
| 3 | 454 (24.3) | 184 (4.7) | 1335 (24.0) |
| Any previous dengue episode (%)‡ | 115 (6.2) | 319 (8.2) | 360 (6.5) |
| Number of past dengue diagnoses (%) |  |  |  |
| 1 | 109 (5.8) | 304 (7.8) | 343 (6.2) |
| ≥2 | 6 (0.3) | 15 (0.4) | 17 (0.3) |
| Diagnostic criteria related to dengue history (%)§ |  |  |  |
| Clinical-epidemiological | 65 (56.5) | 156 (48.9) | 140 (38.9) |
| Laboratory | 35 (30.4) | 145 (45.5) | 198 (55.0) |

\* Median and interquartile range.
† Denominator relative to the total number of women.
‡ Probable cases of dengue reported on Sinan, including cases confirmed by clinical, epidemiological and laboratory criteria, as well as inconclusive cases, with the onset of symptoms between January 2008 and July 2016.
§ The denominator is the number of cases with a history of dengue.

The effectiveness of vaccination on the primary outcome was modified by dengue history, as evidenced by the ratio of ORs (without/with dengue history) in the control groups. With the TN control group, the ratio of ORs was 3.71 (95% CI: 2.15–6.39; p<0.001) for at least one dose and 3.86 (95% CI: 2.0–7.10; p<0.001) for three doses. Similarly, with the OHP control group, the ratio of ORs was 3.40 (95% CI: 2.42–4.79; p<0.001) for at least one dose and 4.11 (95% CI: 2.45–6.88; p<0.001) for three doses.

In the stratum with a history of dengue, receiving at least one dose and the full course of vaccination were both associated with a significant reduction of the primary outcome (total dengue cases), as well as cases of DENV-1 and DENV-2, in the analyses conducted with both control groups. Thus, the VE of the partial and complete vaccination courses was over 70% for total dengue cases, and higher than 60% for the specific serotypes DENV-1 and DENV-2. As there were only 12 cases of DENV-4 with a history of dengue, none of which occurred in vaccinated individuals, the OR was equal to zero (VE=100%), and an adjusted estimate could not be obtained (as non-vaccination perfectly predicted the event).

Additionally, although statistically non-significant, the vaccinated population had a lower incidence of hospitalizations than the non-vaccinated in the stratum with history of dengue (Table 4).

**Table 4.** Association between of vaccination and dengue cases by dengue history and type of control *, 28 municipalities, Paraná, 2019 -2020.

| Outcome | TN control group |  |  |  |  |  | OHP control group |  |  |  |  |  |
| --- | --- | --- | --- | --- | --- | --- | --- | --- | --- | --- | --- | --- |
|  | Vaccinated with at least one dose |  |  | Complete course (3 doses) |  |  | Vaccinated with at least one dose |  |  | Complete course (3 doses) |  |  |
|  | aOR | (95% CI) | VE (%) | aOR | (95% CI) | VE (%) | aOR | (95% CI) | VE (%) | aOR | (95% CI) | VE (%) |
| **With a history of dengue** |  |  |  |  |  |  |  |  |  |  |  |  |
| Dengue | 0.27 | (0.17–0.42) | 73.1 | 0.29 | (0.15–0.57) | 70.6 | 0.25 | (0.17–0.36) | 74.9 | 0.22 | (0.12–0.42) | 78 |
| DENV-1 | 0.32 | (0.16– 0.62) | 68.4 | 0.27 | (0.07–0.99) | 73.1 | 0.27 | (0.15–0.48) | 73.1 | 0.11 | (0.04–0.28) | 89.3 |
| DENV-2 | 0.3 | (0.16–0.54) | 70.4 | 0.34 | (0.15–0.74) | 66.4 | 0.29 | (0.18–0.46) | 71.3 | 0.31 | (0.17–0.56) | 68.8 |
| DENV-4 † | 0 | (0–3.99) | 100 | 0 | (0–11.29) | 100 | 0 | (0–0.4) | 100 | 0 | (0–1.01) | 100 |
| Hospitalizations ‡ | 0.15 | (0.01–1.51) | 85.3 | - | - | - | 0.21 | (0.19–2.27) | 79.5 | - | - | - |
| **No history of dengue** |  |  |  |  |  |  |  |  |  |  |  |  |
| Dengue | 1 | (0.63–1.59) | 0.2 | 1.11 | (0.78–1.60) | -11.2 | 0.85 | (0.65–1.12) | 14.8 | 0.89 | (0.69–1.15) | 11.4 |
| DENV-1 | 0.67 | (0.38–1.18) | 33 | 0.62 | (0.39–0.98) | 38 | 0.57 | (0.44–0.75) | 42.9 | 0.52 | (0.40–0.67) | 48.5 |
| DENV-2 | 1.48 | (1.22–1.82) | -48 | 1.68 | (1.38–2.03) | -67.7 | 1.22 | (1.08–13.8) | -22.1 | 1.27 | (1.07–1.5) | -26.9 |
| DENV-4 § | 0.13 | (0.07–0.24) | 87.1 | 0.23 | (0.10–0.54) | 77.1 | 0.14 | (0.08–0.25) | 86.1 | 0.20 | (0.09–0.45) | 79.7 |
| Hospitalizations | 1.27 | (0.55–2.96) | -27 | 1.49 | (0.62–3.57) | -49 | 1.26 | (0.64–2.49) | -26.8 | 1.41 | (0.65–3.05) | -40.7 |

aOR, odds ratio adjusted for the previous incidence in municipality and, unless otherwise specified, for sex and age. VE, vaccine effectiveness (1- aOR).
\* Unless otherwise specified, the measurements were estimated based on a multilevel analysis with data aggregated at the municipal level and adjusted for the previous municipal incidence of dengue and the individual's sex and age.
† In the group with a history of dengue, this OR was equal to zero because none of the 12 cases of DENV-4 was vaccinated, which prevented obtaining adjusted estimates (as non-vaccination perfectly predicts the event). Therefore, in this stratum, crude ORs and Cornfield exact confidence interval were calculated. However, since this estimate was restricted to two contiguous municipalities (Foz do Iguaçu and Santa Terezinha do Itaipu), where occurred 99% of the cases of DENV-4, we considered the estimated adjusted for previous incidence in municipality (but not for sex and age).
‡ There were four hospitalizations among individuals with a history of dengue (1 vaccinated and 3 unvaccinated). This was not estimable for the full course as there were no cases of hospitalization among individuals with a history of dengue.
§ In the category without a history of dengue, this estimation was restricted to two contiguous municipalities (Foz do Iguaçu and Santa Terezinha do Itaipu), with 99% of the cases of DENV-4 (exempting the need for a multilevel analysis or adjustment for the previous municipal incidence).

For individuals with no history of dengue, vaccination, either at least one dose or the full course, was not associated with the primary outcome. Substantial variations were observed in the association measures according to serotypes. Specifically, for DENV-1, the association was negative (protective) for the three-dose regimen with the two control groups (VE of 38% and 48.5% with the TN and OHP control groups, respectively) and for at least one dose with the OHP group (VE: 42.9%). For DENV-2, the association was positive with both regimens and for both control groups. For DENV-4, a negative association was observed, and effectiveness was more than 70% for both the partial and full vaccination courses, reaching 89.3% for individuals who received the full vaccination course in the evaluation with the OHP control group. In this stratum, no significant association existed between any vaccination regimen and hospitalization for dengue (Table 4).

## DISCUSSION

We used a case-cohort design to estimate the effectiveness of dengue vaccination with at least one dose of CYD-TDV during a campaign in an endemic population. Our findings indicated that vaccination was associated with a 21% reduction in dengue cases in the 15-27 age group across the 28 municipalities in the state of Paraná that participated in the campaign and reported dengue cases in 2019 and 2020. Furthermore, the case-control design suggested that the population-level impact of the vaccination is primarily driven by a reduction in cases among individuals who had previously been exposed to the disease. These results are congruent with those observed in controlled clinical trials, where the protective effect of vaccination was only evident among individuals with serological evidence of previous infection [7].

Conversely, vaccination did not demonstrate a significant advantage in preventing the primary outcome among individuals without a prior dengue history. While vaccination was found to be associated with a decrease in cases of DENV-1 and DENV-4, there was a positive association with cases of DENV-2 in this group. It may be because the vaccine induces immunopotentiation mechanisms that specifically increase the pathogenicity of the DENV-2 serotype [7,19–21].

One of the limitations of the case-control study is that the history of dengue might have been affected by the low sensitivity of the surveillance system for previous events. Nevertheless, it is noteworthy that using the history of a probable case of dengue can assist in identifying a group that could benefit from vaccination. This method of classification based on epidemiological surveillance would be readily implementable in regions where dengue is endemic, particularly in areas where access to laboratory testing for serological status is limited. On the other hand, individuals lacking a history of dengue may not have an initial indication of receiving this dengue vaccine. Therefore, when available, serological testing can be focused on this last population to identify seropositive groups in which the CYD-TDV vaccine is currently recommended.

Another issue is the variation in circulating serotypes, which interferes with the assessment of effectiveness for all cases. In the present study, there were no cases of DENV-3, but there was a high circulation of DENV-2 against which Dengvaxia has lower efficacy.

The estimation of the effectiveness of a vaccine is a complex task due to the difficulty in identifying an appropriate comparison group. Methods to select controls, such as using individuals who have tested negative for the disease or cases of other illnesses, may be prone to biases stemming from patterns of medical consultation and reporting [22,23]. Furthermore, using controls drawn from the same family or neighbourhood as the patients pose the issue of similar vaccination exposure, as access to preventive interventions tends to be similar within these groups [15]. This phenomenon may explain the findings of a recent study conducted in the same region as the present study, in which 618 patients with dengue were compared with 1236 controls composed of neighbours and study or work colleagues [24]. The frequency of vaccination with at least one dose was 43.8% for the controls, which was similar to what was observed in the cases (41.3%), but much lower than that reported for the population in vaccination records (60.5%) [6]. Analyses performed in that study did not weight the controls, leading to a high risk that the control group does not accurately reflect the prevalence of vaccination in the source population, which may explain the failure to identify a significant reduction of dengue cases attributable to the vaccine [24].

In our study, we used official statistics and the vaccination campaign records to recreate a population cohort for comparison purposes. Additionally, by weighting the controls, we aimed to correct or minimize any selection bias [25]. As a result, the weighted controls were more representative of the campaign’s target population regarding critical variables such as vaccination status, sex, age group, and areas. Furthermore, our analysis incorporated adjustments for relevant covariates to control for confounding, which led to results consistent with the efficacy estimated in clinical trials [7].

In conclusion, this study demonstrated that mass vaccination with CYD-TDV significantly reduced the incidence of dengue cases in the campaign’s target population. Additionally, the case-control design suggested that this reduction was primarily driven by the benefit experienced by individuals with a previous history of dengue. We argue that clinical history, as recorded by epidemiological surveillance, could be used as a criterion for recommending CYD-TDV vaccination, particularly in endemic regions with limited access to serological tests.

## Data Availability

All data produced in the present study are available upon reasonable request to the authors.

## Authors’ contributions

FADQ designed the study, analysed the data, and prepared the first draft of the manuscript. DSC participated in the conceptualization of the study, funding acquisition, project administration, data curation and validation. SMR participated in the study conception, data analysis and interpretation of results. SES participated in the conceptualisation, formal analysis, validation and interpretation of results. AMM, MCVCR, LS, MCMB and EMCPM participated in the investigation, validation and interpretation of results. GG participated to the data curation and software use. GA and CP contributed the investigation and validation. KRL participated in the conceptualization of the study, funding acquisition, data curation, validation, data analysis and interpretation of results.

All authors provided relevant input for manuscript writing and review and have read and approved the final manuscript. Thus, the authors sufficiently participated in the work which enables each to take public responsibility for appropriate portions of the content. Therefore, they agree to be accountable for all aspects in ensuring that questions related to the accuracy or integrity of any part of the work are appropriately investigated and resolved.

## SUPPLEMENTARY MATERIALS

**Table S1.** Blocking strategy.

|  |
| --- |
| 1- PBLOCK + UBLOCK + ANONASC + PBLOCK_M + SEX + MUNICIPALITY |
| 2 - PBLOCK + UBLOCK + ANONASC + PBLOCK_M |
| 3 - PBLOCK + ANONASC + PBLOCK_M + SEX |
| 4 - BLOCK + ANONASC + PBLOCK_M + SEX |
| 5 - PBLOCK + UBLOCK + PBLOCK_M + UBLOCK_M |
| 6 - PBLOCK + UBLOCK + ANONASC |
| 7 - PBLOCK + UBLOCK + SEX |

Note: Duplicate records of vaccinated individuals were later excluded. PBLOCK: 'patient's fist name; BLOCK: patient's last name; PBLOCK\_M: 'mother's first name; UBBLOCK\_M: mother's last name.

**Figure S1.**
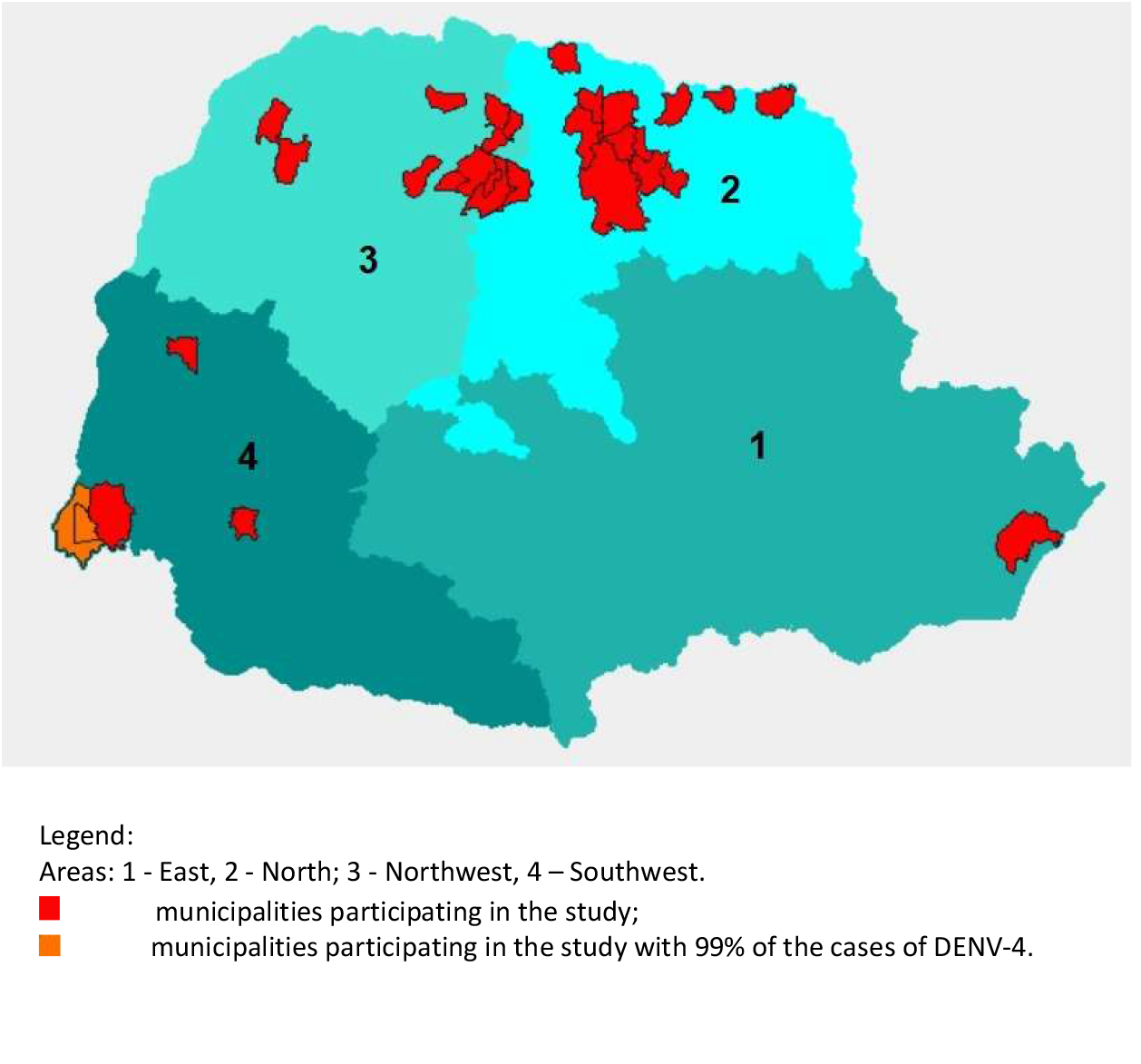
Geographical distribution of areas defined for weighting.

**Table S2.**
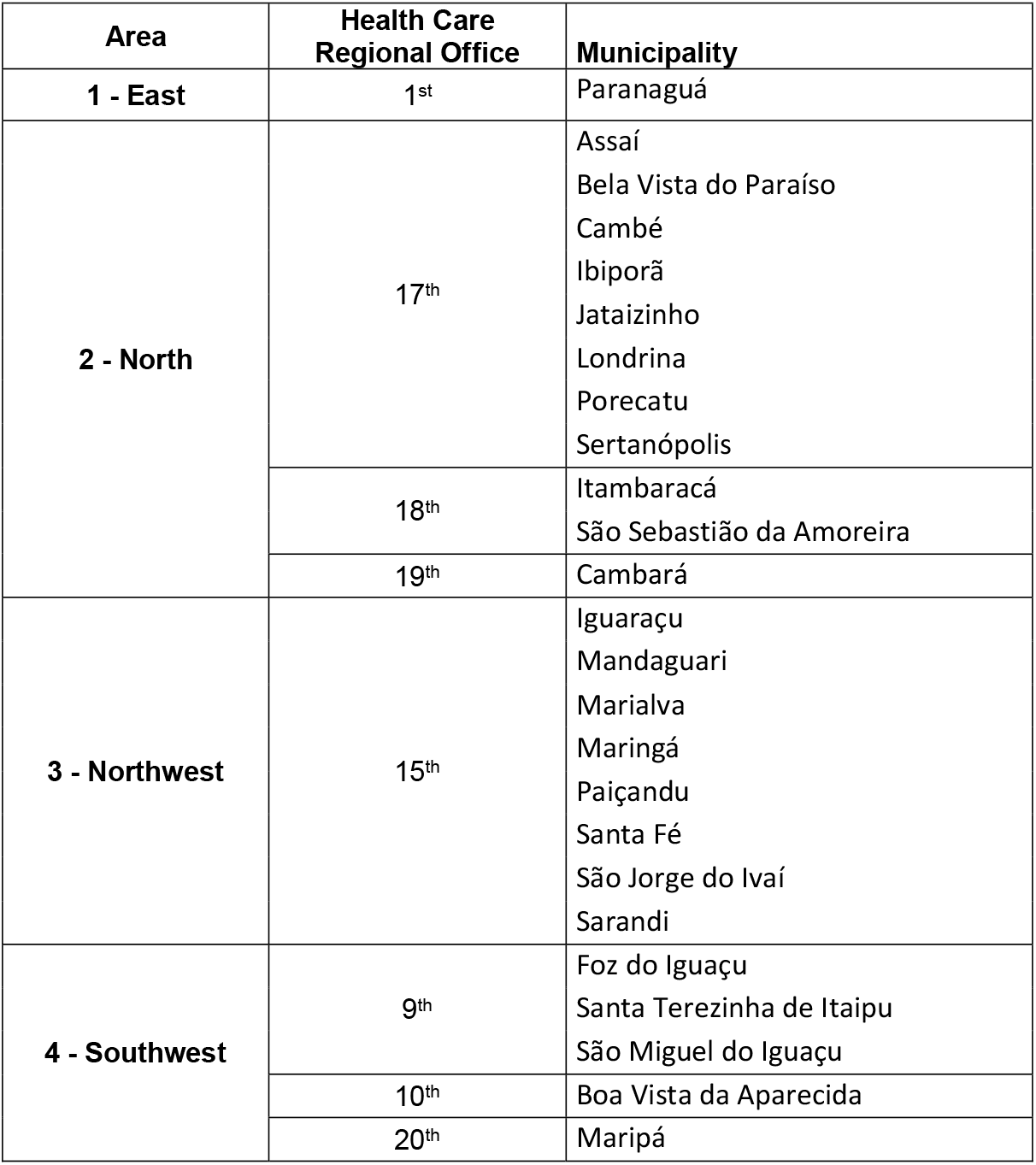
Organization of areas defined for weighting.

**Table S3.** Comparison of adjustment options in case-cohort and case-control designs.

| Characteristics of the logistic model | Outcome | Comparison of cases vs. cohort |  | Case-control designs |  |  |  |  |
| --- | --- | --- | --- | --- | --- | --- | --- | --- |
|  |  | OR | p-value | Weighted | TN controls |  | OHP controls |  |
|  |  |  |  |  | OR | p-value | OR | p-value |
| Ordinary; adjusted for previous incidence, age group, and sex | Dengue | 0.75 (0.68–0.82) | <0.001 | No | 8.91 (7.75–10.24) | <0.001 | 1.08 (0.98–1.21) | 0.13 |
|  |  |  |  | Yes | 0.76 (0.64–0.89) | 0.001 | 0.75 (0.67–0.84) | <0.001 |
|  |  |  |  | Yes* | 0.75 (0.64–0.89) | 0.001 | 0.73 (0.65–0.81) | <0.001 |
| Multilevel; adjusted for age group, and sex | Dengue | 0.79 (0.72–0.87) | <0.001 | No | 8.96 (7.73–10.38) | <0.001 | 1.18 (1.05–1.32) | 0.004 |
|  |  |  |  | Yes | 0.91 (0.58–1.42) | 0.68 | 0.82 (0.62–1.08) | 0.15 |
| Multilevel; adjusted for previous incidence, age group, and sex | Dengue | 0.79 (0.72–0.87) | <0.001 | Yes* | 0.91 (0.58–1.42) | 0.68 | 0.79 (0.6–1.03) | 0.08 |
| Multilevel; adjusted for previous incidence, age group, and sex | Hospitalization for dengue | 1.12 (0.74–1.7) | 0.58 | Yes* | 1.13 (0.49–2.59) | 0.77 | 1.16 (0.58–2.32) | 0.68 |
| Multilevel; adjusted for previous incidence, age group, and sex | DENV-1 | 0.57 (0.48–0.68) | <0.001 | Yes* | 0.64 (0.39–1.06) | 0.08 | 0.55 (0.43–0.7) | <0.001 |
| Multilevel; adjusted by previous incidence, age group, and sex; restricted to municipalities with DENV-1 | DENV-1 | 0.58 (0.48–0.68) | <0.001 | Yes* | 0.64 (0.39–1.07) | 0.09 | 0.55 (0.43–0.71) | <0.001 |
| Multilevel; adjusted for previous incidence, age group, and sex | DENV-2 | 1.08 (0.95–1.22) | 0.23 | Yes* | 1.31 (1.07–1.61) | 0.008 | 1.1 (0.97–1.25) | 0.14 |
| Multilevel; adjusted by previous incidence, age group, and sex; restricted to municipalities with DENV-2 | DENV-2 | 1.08 (0.96–1.22) | 0.22 | Yes* | 1.31 (1.07–1.61) | 0.008 | 1.1 (0.97–1.25) | 0.14 |
| Ordinary; adjusted for previous incidence, age group, and sex; restricted to municipalities with DENV-4 | DENV-4 | 0.13 (0.07–0.23) | <0.001 | Yes* | 0.11 (0.06–0.2) | <0.001 | 0.11 (0.06–0.2) | <0.001 |

\* Age was included as a quantitative variable (in years) in these case-control analyses.

